# Using Blood-Test Parameters to Define Biological Age among Older Adults: Association with Morbidity and Mortality Independent of Chronological Age Validated in Two Separate Birth Cohorts

**DOI:** 10.1101/2022.02.12.22270832

**Authors:** Johanna Drewelies, Gizem Hueluer, Sandra Duezel, Valentin Max Vetter, Graham Pawelec, Elisabeth Steinhagen-Thiessen, Gert G. Wagner, Ulman Lindenberger, Christina M. Lill, Lars Bertram, Denis Gerstorf, Ilja Demuth

## Abstract

Biomarkers defining biological age are typically laborious or expensive to assess. Instead, in the current study, we identified parameters based on standard laboratory blood tests across metabolic, cardiovascular, inflammatory, and kidney functioning that had been assessed in the Berlin Aging Study (BASE; *n* = 384) and Berlin Aging Study II (BASE-II, *n* = 1,517). We calculated biological age using those 12 parameters that individually predicted mortality hazards over 26 years in BASE.

In BASE, older biological age was associated with more physician-observed morbidity and higher mortality hazards, over and above the effects of chronological age, sex, and education. Similarly, in BASE-II, biological age was associated with physician-observed morbidity and subjective health, over and above the effects of chronological age, sex, and education as well as alternative biomarkers including telomere length, DNA methylation age, skin age, and subjective age. We discuss the importance of biological age as one indicator of aging.

## Introduction

It is an everyday observation that some people seem to be significantly younger in their physical appearance and behavior than one would expect based on how old they are chronologically, whereas others seem to be much older than they are chronologically. In other words, one of the hallmarks of old age is its high degree of heterogeneity between people, even if these people are of the same chronological age ^1,2^. This heterogeneity is known to increase with age ^3^. It is thus desirable to move beyond chronological age as a proxy for underlying biological aging processes and to use biological markers available that allow quantifying people’s risk of developing age-associated deficits and diseases. One rather conservative definition of such a marker of biological age has been proposed by the American Federation for Aging Research (AFAR) stating that biological age must be able to predict the rate of aging and it must be a better predictor of life span than chronological age ^4–6^. In addition, biological age must have the capability to monitor one or more basic processes that contribute to or underlie aging rather than merely representing effects of disease, be tested without harming the person, and work in humans and laboratory animals alike ^5,7^.

To date, however, no such marker fulfilling the AFAR defining features has been identified. As a consequence, a number of alternative definitions have been proposed ^5,7^. One of the most established alternatives was formulated by was proposed by Jylhäva and colleagues ^3^ who suggest that a marker of biological age must predict existing and prospective age-associated phenotypes over and above chronological age. To this end, several single measures of biological age have been examined. To illustrate, a well-established biological aging marker is represented by the length of chromosome end repeats, i.e., the telomeres. These chromosomal regions reflect the replicative age of a given cell, such that the average of the telomere lengths of a cell or tissue sample is weakly associated with chronological age as well as with a multitude of aging phenotypes ^8–11^. Relative leukocyte telomere length has been assessed in the Berlin Aging Study II (BASE-II), one of the two cohorts studied here, and similar to other groups, we delineated associations with age-related phenotypes and lifestyle ^12–15^. As another example biomarker is the epigenetic clock variable, which aims to measure DNA methylation (DNAm) age and has recently been described to be moderately to strongly associated with chronological age ^15–18^. Additionally, DNAm age and its deviation from chronological age, the DNAm age acceleration, have been suggested to reflect biological age ^16^. Interestingly, DNA methylation age measures appear not to be related to telomere lengths and therefore might reflect different aspects of biological ageing ^17,19–23^.

While these two sets of measures have the potential to quantify certain aspects of biological aging, they are likely unable to capture the aging process as a whole. As a consequence, other approaches to estimate biological age by combining several individual biomarkers have been proposed^24^ and these algorithms, too, differ in their accuracy to quantify biological aging. For instance, by comparing five different biological age algorithms, Levine has shown that the algorithm by Klemera and Doubal, which combines 10 different biomarkers performed best in the National Health and Nutrition Examination Survey III (NHANES-III, N=9,389) cohort in predicting mortality hazards ^25^. Employing this algorithm ^26^ and parameters estimated from the NHANES-III dataset, and considering the exact same 10 biomarkers used in NHANES-III, Belsky and colleagues calculated biological age in the Dunedin study ^27^. Biological age was estimated for this birth cohort at age 38 years and then used to calculate the rate of aging based on longitudinal data on 18 biomarkers reflecting different areas of health (e.g., cardiovascular, metabolic). Their results indicated that individuals with evidence for decelerated biological aging showed better physical health and cognitive performance ^27^. Instead of mortality-predicting biomarkers, Sebastiani and colleagues used 19 age-associated parameters to identify 26 different biomarker signatures in the Long Life Family Study. Ten of these signatures were then found to be associated with mortality hazards, morbidity, and physical functioning, and seven of these signatures could be replicated in an independent cohort ^28^.

Importantly, previous studies have found at best moderate correlations between the various biological age measures, such as telomere length, epigenetic clocks, or biomarker composites ^17,20,29,30^ suggesting that even though these measures tap into the same overarching construct space, they still capture different and unique aspects of aging processes. One potential weakness of the biomarkers included in previous composite scores is that they were typically selected based on their association with chronological age. ^20,27^ In addition, for practical purposes, some of these biomarkers are laborious or expensive to determine.

In the current study, we therefore opted for an alternative strategy and operationally defined a biological age composite using standard validated clinical laboratory blood tests across metabolic, cardiovascular, inflammatory, and kidney functioning with respect to their association with mortality. This allowed us to capture the multidimensionality and multifunctionality of biological age beyond single measure age indicators. To identify relevant laboratory blood parameters, we made use of a second and independent study of older adults for whom mortality information was available. Laboratory blood parameters that were identified as mortality-relevant were then used to calculate a biological age composite. Thus, in the present study we aimed to identify parameters based on standard laboratory blood tests across multiple domains of functioning in two independent data sets and examined whether and how individual differences in biological age are associated with sociodemographic and health measures that may have operated as antecedents, correlates, or outcomes.

## Results

### Variables Defining Biological Age

In a preliminary step, we used mortality information available for the earlier-born cohort of participants in the Berlin Aging Study (BASE) and estimated a series of 33 separate Cox proportional hazards regression models. ^31^ We used these to identify mortality-relevant relevant blood laboratory parameters markers reflecting metabolic, cardiovascular, inflammatory, and kidney functioning that had been measures in both BASE study and its successor study, BASE-II. As shown in the Appendix, we identified 12 parameters that were predictive of mortality hazards in BASE (see Appendix Table A.1), whereas 21 parameters were not predictive of mortality (see Appendix Table A.2). Descriptive statistics and intercorrelations among the 12 mortality-relevant parameters that were subsequently used to calculate the biological age composite are presented in Table 1. Intercorrelations are generally in the small to moderate range, with the single largest intercorrelation being r = .50_BASE_ (between leukocytes and lymphocytes in BASE) and r = .61_BASE-II_ (between leukocytes and lymphocytes in BASE-II). These divergences suggest that all these parameters represent different aspects of biological aging processes.

**Table 1.** Means, Standard Deviations, and *Intercorrelations among the Variables that constitute the Biological Aging Composite Measures in the Berlin Aging Study (below the Diagonal) and the Berlin Aging Study II (above the Diagonal)*

|  | Intercorrelations |  |  |  |  |  |  |  |  |  |  |  |
| --- | --- | --- | --- | --- | --- | --- | --- | --- | --- | --- | --- | --- |
|  | 1 | 2 | 3 | 4 | 5 | 6 | 7 | 8 | 9 | 10 | 11 | 12 |
| $M_{BASE-II}$ | 12.43 | 139.57 | 102.10 | 5.34 | 62.24 | 8.76 | 3.47 | 5.60 | 13.85 | 5.81 | 1.70 | 68.84 |
| $SD_{BASE-II}$ | 1.82 | 2.77 | 2.90 | 1.34 | 3.23 | 1.47 | 0.61 | 0.51 | 1.13 | 2.08 | 1.24 | 11.10 |
| (1) Zinc (4.70–17.80) | 1 | **.11** | -.01 | .02 | .05 | -.01 | -.06 | .05 | .17 | **-.01** | -.01 | .01 |
| (2) Sodium (131–150) | **.14** | 1 | **.61** | **.01** | .14 | **-.11** | **-.09** | **-.11** | **.05** | -.04 | -.03 | -.04 |
| (3) Chloride (92–122) | .02 | **.45** | 1 | -.03 | **.08** | **-.05** | -.03 | **-.14** | -.03 | -.04 | .00 | -.02 |
| (4) Uric acid (1.50–12.50) | **-.14** | -.09 | **-.13** | 1 | **-.17** | **.12** | .06 | **.18** | **.31** | **.18** | **.10** | **-.32** |
| (5) Albumin (42.80–72.50) | **.31** | **.10** | .08 | **-.19** | 1 | **-.50** | **-.40** | **-.23** | .05 | **-.10** | .02 | .03 |
| (6) Alpha-1 globulin (1.90–5.30) | -.08 | .08 | .05 | -.00 | **-.28** | 1 | **.61** | **.29** | -.04 | .17 | .03 | -.02 |
| (7) Alpha-2 globulin (5.00–16.80) | -.05 | -.02 | -.02 | **.13** | **-.41** | **.34** | 1 | **.12** | -.01 | .12 | **.03** | .00 |
| (8) HbA1c (3.90–12.10) | -.02 | **-.22** | **-.16** | **.11** | -.06 | -.01 | **.20** | 1 | -.02 | **.14** | **.08** | -.05 |
| (9) Hemoglobin (7.00–18.50) | **.25** | .05 | -.05 | -.00 | **.17** | -.08 | -.10 | .02 | 1 | **.06** | .01 | .03 |
| (10) Leukocytes (2.10–13.00) | -.04 | -.00 | -.05 | **.13** | **-.15** | **.16** | **.18** | **.11** | **.17** | 1 | **.78** | **-.10** |
| (11) Lymphocytes (0.30–5.00) | .09 | .04 | .02 | .01 | .02 | -.05 | -.02 | .08 | **.20** | **.50** | 1 | -.04 |
| (12) Creatinine (13.86–131.60) | **.12** | .01 | .05 | **-.43** | **.14** | .09 | **-.21** | **-.13** | **.29** | -.03 | **.11** | 1 |
| $M_{BASE}$ | 11.37 | 141.18 | 104.27 | 5.45 | 57.65 | 3.55 | 9.54 | 6.28 | 14.00 | 6.63 | 1.71 | 52.18 |
| $SD_{BASE}$ | 1.98 | 2.78 | 3.91 | 1.53 | 4.37 | 0.61 | 1.56 | 1.23 | 1.46 | 1.82 | 0.64 | 16.26 |

Note. $N = 384$ . Intercorrelations in bold differ statistically significantly from zero at $p < .05$ .

### Biological Age Composite in BASE

In the next step, we calculated the biological age composite for BASE participants. The upper portion of Table 2 presents the descriptive statistics of chronological age, biological age, and the difference between biological age and chronological age. It can be seen that, as a direct consequence of the procedures applied, the average biological age (84.35 years) was the same as the average chronological age (84.35 years). Most important for the research question of this study are the individual differences between a given participant’s biological age and chronological age. The individual distribution of biological age by chronological age is graphically shown in the left-hand portion of Figure 1, separately for men and women. These data indicate that among participants aged 70 years and older, the person with the youngest biological age is equivalent to 40 years and the person with the highest biological age is 119 years. Likewise, the discrepancy between biological age and chronological age ranges between – 34 (indicating that, biologically, this person is 34 years younger than one would expect based on his or her chronological age) and 27 (indicating an age acceleration of 27 years when comparing biological to chronological age). Of note is also that the correlation between chronological age and biological age is *r* = .65 among women and *r* = .70 among men, indicating that participants who are older chronologically also tend to be older biologically, but that there are also individual differences in how chronological age and biological age coincide.

**Table 2.** Descriptive Statistics for Chronological Age, Biological Age, and the Difference between Biological Age and Chronological Age, separately for the Berlin Aging Study and the Berlin Aging Study II

|  | *N* | *M* | *SD* | *Min–Max* |
| --- | --- | --- | --- | --- |
| Berlin Aging Study |  |  |  |  |
| Chronological age | 384 | 84.35 | 8.74 | 70 – 102 |
| Biological age | 384 | 84.35 | 12.70 | 40 – 119 |
| $\Delta$ biological age – chronological age | 384 | 0 | 9.36 | –34 – 27 |
| Berlin Aging Study II |  |  |  |  |
| Chronological age | 1,517 | 68.66 | 3.62 | 60 – 85 |
| Biological age | 1,517 | 63.48 | 8.33 | 37 – 90 |
| $\Delta$ biological age – chronological age | 1,517 | –5.19 | 7.86 | –29 – 23 |

**Figure 1.**
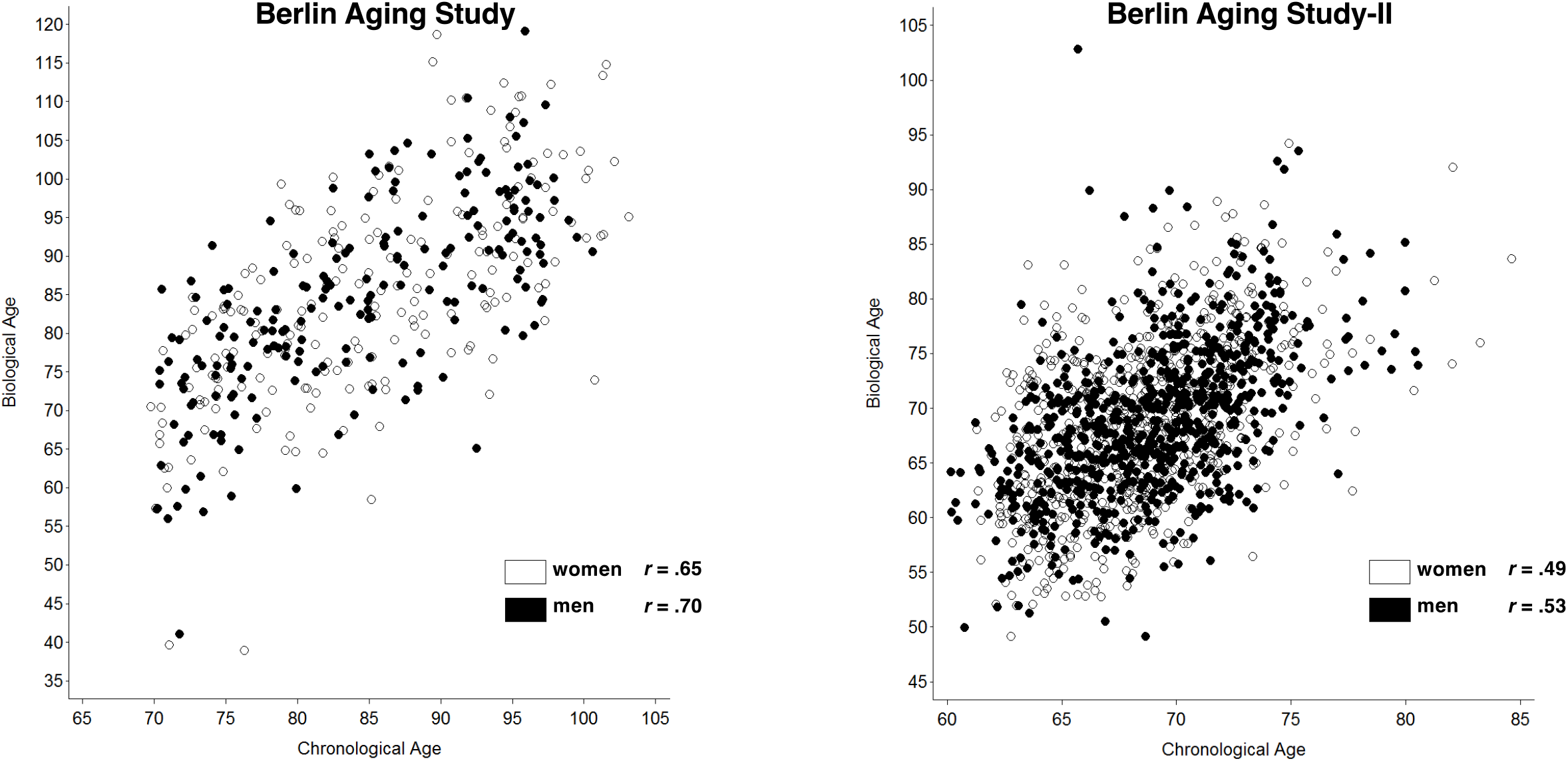
Biological age in the Berlin Aging Study (left-hand panel) and the Berlin Aging Study II (right-hand panel).

Table 3 reports results of Cox proportional hazards regression models testing the predictive performance of biological age and the socio-demographic correlates for mortality hazards. As one would expect, being older was associated with greater mortality risk (Hazard Ratio, HR = 1.08, 95% Confidence Interval, CI: 1.06 – 1.10) and being a woman was independently associated with lower mortality risk (HR = 0.72, 95% CI: 0.58 – 0.88, both *p*’s < .05). There was no effect of socio-economic status (SES) in this population (*p* > .10). Most important for our research question is that biological age exhibits additional unique predictive effects for mortality hazards over and above those found for chronological age and sex (HR = 1.03, 95% CI: 1.02 – 1.04, *p* < .05). With every one additional year of biological age, the residualized risk of death increased by 3%. Figure 2 shows the Kaplan-Meier survival curves for two groups above or below the median biological age. Those who are biologically younger relative to their chronological age live longer than those who are biologically older relative to their chronological age, a difference in average survival probability that amounts to more than 1.5 years.

**Table 3.** Hazard Ratios for Mortality by Biological Age and the Correlates in the Berlin Aging Study

| Predictors | Hazard Ratio | [95% CI] |
| --- | --- | --- |
| Chronological age | 1.08* | [1.06–1.10] |
| Women | 0.72* | [0.58–0.88] |
| Education | 0.99 | [0.94–1.04] |
| Biological age | 1.03* | [1.02–1.04] |

*Note.* $N = 384$ .
\* $p < .01$ .

**Figure 2.**
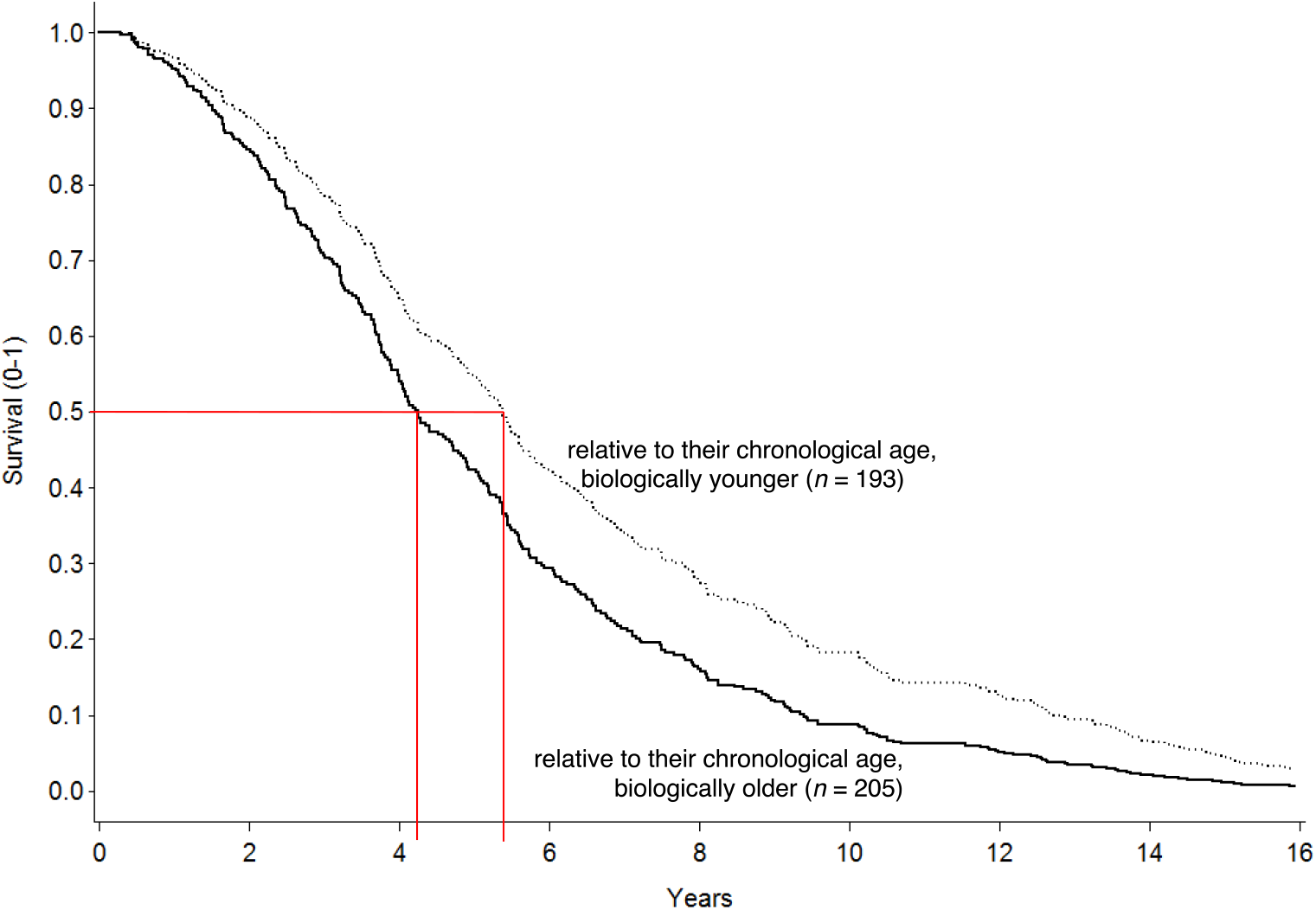
Kaplan-Meier survival curves in the Berlin Aging Study for two groups based on a median cutoff for biological age. Those who are biologically younger relative to their chronological age live longer than those who are biologically older relative to their chronological age, a difference in average survival probability that amounts to more than 1.5 years. Curves are residualized for differences by age, sex, and education.

In a series of follow-up analyses, we tested the unique predictive effects for mortality hazards of biological age compared to those of chronological age, sex, and SES. Results are summarized in the Appendix (Table A.3). From the middle column, it can be seen that chronological age, sex, SES, and biological age together accounted for 26.81% of the variance in mortality hazards among BASE participants. From the right-hand column, it is clear that, as one would expect, chronological age yields the largest unique predictive effects in this age range, whereas the effects of sex and SES were negligible. In contrast, biological age had a unique predictive impact on mortality hazards that (a) was over and above that of chronological age, (b) was more than 2.5-fold the effect of sex, and (c) was smaller than the effect of chronological age. The majority of the variance is shared between predictors (8.21 + 0.77 + 0.02 + 2.17 = 11.17 unique variance out of 26.81 total variance amounts to 42% unique variance and 58% shared variance).

### Correlates of Biological Age in BASE

In a third step, we used regression analyses to examine whether and how socio-demographic and physical health predict individual differences in biological age. Results are presented in the middle-column of Table 4 and graphically illustrated in the left-hand panel of Figure 3. It can be seen that being older chronologically was associated with being older biologically (*β*= .638, *p* < .05). Important for the question at hand, morbidity (*β*= .114, *p* < .05) was also uniquely associated with biological age, over and above the effects of chronological age, sex, and SES. Participants who suffer from more physical illnesses (see left-hand portion of Figure 3) were more likely to be biologically older than they were chronologically relative to their peers with fewer diseases.

**Table 4.** Predicting Biological Age from Chronological Age, Sex, Education, and Physician-Observed Morbidity, separately for the Berlin Aging Study and the Berlin Aging Study-II

| Predictors | Dependent variable: Biological age |  |
| --- | --- | --- |
|  | Berlin Aging Study | Berlin Aging Study II |
| Chronological age | .638* | .504* |
| Women | -.020 | -.001 |
| Education | .036 | -.028 |
| Morbidity | .114* | .108* |
| Total $R^2$ | .448 | .274 |
| $F(df1, df2)$ | 69.08* (4, 345) | 113.55* (4, 1,205) |

*Note.* Standardized regression coefficients are reported. Berlin Aging Study: $N = 397$ . Berlin Aging Study: $N = 1,210$ . Age and women centered at sample mean, all other predictors z-standardized.
<sup>a</sup> $p = .05$ . \* $p < .05$ .

**Figure 3.**
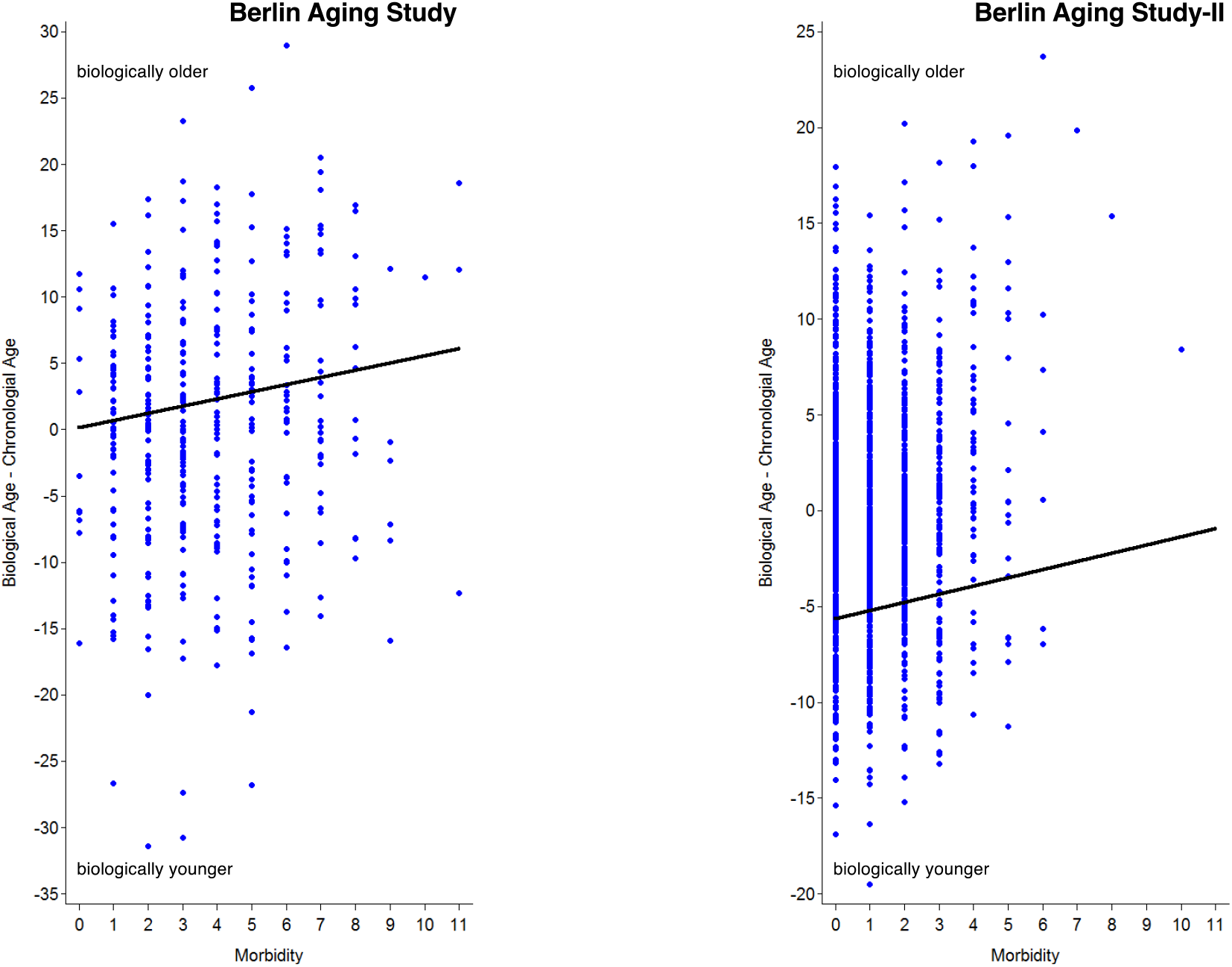
Linking physician-observed morbidity with biological age in the Berlin Aging Study (left-hand panel) and the Berlin Aging Study-II (right-hand panel). Consistent across sample, it can be seen that participants who had more physical illnesses were more likely to be biologically older than they were chronologically, as compared with their peers with fewer diseases.

### Biological Age Composite in BASE-II

In the next step, we used the independently identified parameters from BASE to calculate a biological age composite in participants of its successor study, BASE-II. Results of this step are reported in Table 2 and the right-hand panel of Figure 1. Three findings are noteworthy. First, on average, BASE-II participants were 6.10 years younger biologically than their chronological age. Second, again we found considerable individual differences in biological age. Among participants chronologically aged 60 years and older, the person with the youngest biological age was 37 years and the person with the oldest biological age was 90 years. Likewise, the discrepancy between biological age and chronological age ranges between –29 (indicating a 29 years younger biological than chronological age) and 23 (indicating a 23 years older biological than chronological age). Third, the correlation between chronological age and biological age is *r* = .49 among women and *r* = .53 among men meaning that, participants who are older chronologically also tend to be older biologically, but there are also individual differences in how chronological age and biological age coincide.

### Correlates of Biological Age in BASE-II

In a final set of analyses, we again examined correlates of biological age. Mimicking our analyses in BASE, we started with regression analyses that link socio-demographic and physical health variables with biological age. Results are presented in the right-hand column of Table 4 and graphically illustrated in the right-hand panel of Figure 3. Similar to BASE, older chronological age was associated with older biological age (*β*= .504, *p* < .05). Importantly, for again, morbidity (*β*= .108, *p* < .05) was uniquely associated with biological age, over and above the effects of chronological age, sex, and SES. Those who suffered from more physical illnesses (see right-hand portion of Figure 3) tended to be biologically older than they were chronologically.

In the last step, we made use of the rich interdisciplinary data in BASE-II that allowed us to examine the role of biological age for various physical health measures while accounting for sociodemographic factors and a number of alternative age biomarkers, including telomere length, DNA methylation age, skin age, and subjective age (intercorrelations are reported in the Appendix Table A.4). Results are reported in Table 5. Over and above the well-known effects of chronological age, sex, and education as well as the alternative age biomarkers, older biological age as assessed by our novel the biological age composite was found to predict higher physician-observed morbidity (*β*= .144, *p* < .05) and lower subjective health (*β*= –.099, *p* < .05), whereas no associations were found with lung capacity (*β*= .004, *p* > .10).

**Table 5.** Predicting Physician-Observed Morbidity, Lung Capacity, and Subjective Health from Five Different Alternative Age Indices: Shared and Unique Effects.

| Predictors | Model 1 | Model 2 | Model 3 | Model 4 | Model 5 | Conjoint model |
| --- | --- | --- | --- | --- | --- | --- |
| **Physician-observed morbidity** |  |  |  |  |  |  |
| Chronological age | .007 | .002 | -.039 | .066* | -.009 | -.064 |
| Women | -.068* | -.053 | -.069* | -.058 | -.034 | -.076 |
| Education | -.018 | -.030 | -.032 | -.029 | -.002 | .014 |
| Telomere length | -.029 | — | — | — | — | -.001 |
| DNA meth<sup>1</sup> | — | -.058 | — | — | — | .040 |
| **Biological age** | — | — | **.145\*** | — | — | **.144\*** |
| Skin age | — | — | — | -.014 | — | .029 |
| Subjective age | — | — | — | — | -.048 | .032 |
| Total $R^2$ | .005 | .008 | .023 | .009 | .003 | .028 |
| $F$ | 1.35 | 2.31 | 7.07* | 2.30 | 0.89 | 2.50* |
| ($df1, df2$) | (4, 1,164) | (4, 1,172) | (4, 1,205) | (4, 987) | (4, 1,127) | (8, 687) |
| **Lung capacity** |  |  |  |  |  |  |
| Chronological age | -.210* | -.220* | -.191* | -.220* | -.223* | -.212* |
| Women | -.710* | -.704* | -.709* | -.715* | -.715* | -.743* |
| Education | .033 | .044 | .047 | .050 | .021 | .039 |
| Telomere length | -.004 | — | — | — | — | -.032 |
| DNA meth<sup>1</sup> | — | .003 | — | — | — | -.045 |
| **Biological age** | — | — | **-.023** | — | — | **.004** |
| Skin age | — | — | — | .005 | — | .010 |
| Subjective age | — | — | — | — | -.018 | .030 |
| Total $R^2$ | .544 | .548 | .556 | .554 | .556 | .559 |
| $F$ | 175.34* | 179.66* | 188.46* | 144.54* | 179.58* | 53.60* |
| ($df1, df2$) | (4, 587) | (4, 594) | (4, 603) | (4, 466) | (4, 573) | (8, 339) |
| **Subjective health** |  |  |  |  |  |  |
| Chronological age | -.025 | -.026 | -.034 | -.063* | .137* | .187* |
| Women | -.090* | -.086* | -.055* | -.033 | -.072* | -.048 |
| Education | -.000 | -.020 | .014 | .022 | -.009 | .019 |
| Telomere length | -.019 | — | — | — | — | -.053 |
| DNA meth<sup>1</sup> | — | .001 | — | — | — | -.003 |
| **Biological age** | — | — | **-.101\*** | — | — | **-.099\*** |
| Skin age | — | — | — | .076* | — | .039 |
| Subjective age | — | — | — | — | -.289* | -.283* |
| Total $R^2$ | .008 | .008 | .011 | .009 | .066 | .066 |
| $F$ | 2.42* | 2.41* | 3.72* | 2.47* | 21.59* | 6.62* |
| ($df1, df2$) | (4, 1,107) | (4, 1,111) | (4, 1,125) | (4, 893) | (4, 1,060) | (8, 645) |

Note. DNA meth = DNA methylation age. standardized prediction effects ( $\beta$ ). <sup>a</sup> $p = .07$ , <sup>b</sup> $p = .05$ , \* $p < .05$ .

## Discussion

In the current study, we identified 12 standard laboratory parameters reflecting metabolic, cardiovascular, inflammatory, and kidney functioning that were each identified as being mortality relevant that were available in two independent cohorts (BASE and BASE-II). These 12 parameters were selected because they showed the strongest association with mortality in the earlier-born cohort (BASE). These mortality-relevant parameters were then used to define a comprehensive multi-indicator biological age composite which was also associated with mortality independently of chronological age. Proceeding to validate biological age in BASE-II as well, we found that in both cohorts biological age was associated with a composite index of morbidity. Because our morbidity index represents a broad range of common diseases (such as diabetes or cancer), we interpret this finding to indicate that our biological age composite also adequately is also a measure of morbidity burden. The associations found with subjective health suggest that our biological age composite is also intertwined with and probably shapes experiential and evaluative factors.

The correlation between chronological age and the biological age composite was lower in BASE-II (*r* = .53 in men and *r* = .49 in women) than in BASE (*r* = .70 in men and *r* = .65 in women). These differences between the two cohorts could be explained by a combination of several factors. First, the BASE-II cohort investigated here (68.8 ±3.7 years) is on average younger than the BASE participants (84.9 ±8.7 years). In addition, the overall BASE-II sample is on average more positively selected on variables such as education, self-reported health and other characteristics, which is related to its recruitment as a convenience sample ^32^, whereas ascertainment of BASE participants was based on city registry information in the aim to collect a representative sample. Another factor in this context is that BASE-II participants were born on average more than 20 years later than participants of the BASE study and thus represent different birth cohorts. We have shown earlier for propensity-score-matched subsamples (matched for age, sex and education) of these two cohorts that the later-born BASE-II participants performed significantly better on cognitive, psychosocial, and physical health variables when compared with same-aged BASE participants born and tested 20 years earlier ^33–35^. Analyzing matched samples was not an option for the current report because this would have reduced the effective sample size to less than n=100 in each cohort. Since the biological age composite relies on laboratory parameters associated with mortality in BASE which was significantly older than BASE-II at baseline, the lower degree of correlation between the biological age composite and chronological age in BASE-II is not unexpected.

BASE was initiated in 1990 and therefore certain laboratory parameters widely available now were not available for this cohort. This is a limitation with respect to the strategy chosen here, where we selected only parameters that were available in both data sets. At the same time, this can also be considered a strength of the current study because the selected parameters today are regularly available as clinical routine parameters for people of the age group investigated here. In other words, the biological age composite can be calculated and might add to the overall view in clinical decision-making without additional costs to assess more sophisticated biological age indicators such as those that consider parameters not routinely clinically available ^20,27^, biomarker signatures ^28^, or metabolic profiles^36^. Another strength is the strategy of drawing on the older BASE study to construct the biological age composite and then evaluate it in the BASE-II cohort. Because several investigators of BASE were also involved in initiating BASE-II, this ensured a high degree of consistency in data collection with respect to the tests employed in the two cohorts investigated. Additionally, participants from both studies were recruited from the same geographical area, Berlin, Germany, which further increases comparability between the two cohorts studied here. Thus, using two independent samples allowed us to cross-validate findings within this study report.

We also note that several approaches have been applied to index biological age which could result in distinct biological age measures ^27^. It is thus crucial to also consider the analytic aspects when examining composite scores of biological age. For instance, in the present study, we calculate the biological age composite following procedures described in Belsky et al. ^27^. It is thus unclear how our biological age measure generalizes beyond the findings obtained here ^37^.

In the present study, we selected a specific set of biomarkers based on availability in both studies and their association with mortality in the earlier BASE study. We are aware that the biological age composite is always dependent on the set of biomarkers chosen. Based on the extant literature, it is unlikely that a study will be able to find an indicator that fulfills all AFAR criteria at the same time. But this does not mean that subsets of biomarkers cannot be used to examine biological age composites. For example, we note that the highest correlation was found between lymphocytes and leukocytes. This is not surprising given that lymphocytes are a subgroup of leukocytes. However, both parameters contribute independently and significantly to the biological age composite. It is therefore necessary to further examine the specific dynamic mechanisms of each biological age component, ideally over time, to better understand how, first, they change over time, and and also how their effect size with respect to contributing to biological age changes over time.

Lastly, our study examined the nature and correlates of biological age in two independent cohorts. Importantly, the correlational design of the cross-sectional data used does not allow us to draw conclusions about the causal or directional mechanisms underlying these associations. For instance, it is conceivable that the predictive validity of individual biomarkers of aging varies across the lifespan, with some being more important at younger ages and others being more important at old ag. Based on the currently available data were are unable to quantify to what extent biological age is related to individual differences in the rate of aging over the course of the second half of life^38^, or whether it could also be a matter of persistent individual differences that have already emerged at earlier stages of life ^39^. Ideally this question should be examined in appropriately designed longitudinal studies. Similarly, even though biological age might be a promising tool as a biomarker to predict diseases before they manifest clinically, and to monitor effects of interventions, the underlying molecular and cellular mechanisms of the observed correlations still remain largely unknown.

## Conclusions

In the present study, we operationally defined biological age as a comprehensive multi-indicator biomarker developed on the basis of a total of 12 single markers reflecting metabolic, cardiovascular, inflammatory, and kidney functioning that had each been identified as being mortality relevant over a >25 year observation period (BASE). In that study, older biological age was associated with more physician-observed morbidity and higher mortality hazards, over and above the well-known effects of chronological age, sex, and education. In the later BASE-II cohort, older biological age was also associated with more physician-observed morbidity and lower subjective health (but not lung capacity), over and above the well-known effects of chronological age, sex, and education as well, even when alternative more recent age biomarkers including telomere length, DNA methylation age, skin age, and subjective age were additionally considered. Our findings suggest that our biological age composite based on convenient and non-invasive, inexpensive, standard validated laboratory blood tests promises to provide unique insights into the heterogeneity of aging and how biological aging processes are intertwined with morbidity, subjective health, and mortality.

## Online Methods

In this report, we used data from the Berlin Aging Study (BASE, obtained 1990–93) and the Berlin Aging Study II (BASE-II, obtained 2010–14). Detailed descriptions of participants, variables, and procedures can be found in previous publications (BASE^40^, BASE-II^32^). Selected details relevant to this report are given below.

### Participants and Procedure

#### Berlin Aging Study (BASE)

The initial BASE sample consisted of 516 residents of former West-Berlin districts (age: *M* = 84.35, *SD* = 8.66, range = 70–103; 50% women) identified based on the obligatory city registry, recruited and tested from 1990–93. Here, we included data from the 384 participants (age: *M* = 84.35, *SD* = 8.59, range = 70–102; 49% women) who had provided information on all variables of interest (see below). Assessments took place either at the hospital (as part of a medical evaluation) or at participants’ places of residence (i.e., private household or institution, for the self-report questionnaires) and were obtained in individual face-to-face sessions by trained research assistants. Sessions required an average of 90 minutes and, when necessary, were split into shorter units of assessment. The Ethics Commission of the Berlin Chamber of Physicians (Ärztekammer Berlin) approved the BASE study prior to the first assessments in 1990 (approvals were not numbered at that time).

#### Berlin Aging Study II (BASE-II)

The BASE-II sample included residents of the greater metropolitan area of Berlin, recruited via a participant pool at the Max Planck Institute for Human Development (Berlin) and via advertisements in local newspapers and the public transportation system. In the current study, we included data from 1,517 older participants (age: *M* = 68.66, *SD* = 3.62, range = 60–85; 51% women) recruited and tested from 2010-2014 and who had provided information on all variables of interest (see below). Measures were obtained either at the Charité University hospital (as part of a medical evaluation) or via self-administered take-home questionnaires. The Ethics Committee of the Charité University hospital approved the BASE-II study (approval number EA2/029/09).

### Measures

#### Biological age

To operationally define biological age, we selected the following 12 standard blood laboratory parameters from a total of 33 parameters across metabolic, cardiovascular, inflammatory, and kidney functioning: zinc, sodium, chloride, uric acid, albumin, alpha-1 globulin, alpha-2 globulin, HbA1c, hemoglobin, leukocytes, lymphocytes, and creatinine. All parameters were measured in a clinical routine laboratory with appropriate quality standards.

#### Correlates

We linked the biological age indices to a number of sociodemographic and health measures that may have served as antecedents, correlates, or outcomes. As far as this was possible for studies conducted some 20 years apart, we used operational definitions of our major constructs that closely mirrored one another.

*Chronological Age* was calculated based on year and month of the assessment/interview relative to the year and month of birth. *Sex* was coded as a binary variable that contrasted women (1) with men (0). Following usual practice in BASE ^41^, *socio-economic status* (SES) was operationally defined using a unit-weighted composite of three measures: (a) equivalent income, defined as the net household income weighted by the number of people sharing the household; (b) occupational prestige, based on a standard rating scale for Germany; and (c) number of years of education (for details, see ^40^). For BASE-II, we also followed the usual practice ^42^and operationally defined socio-economic status as the years of education received.

In both studies, *morbidity* was assessed as part of the medical examinations carried out by physicians. Diagnoses were obtained through participant reports, with certain diagnoses (e.g., diabetes mellitus) being verified by additional (blood-laboratory) tests. For BASE, we used the number of physician-observed diagnoses of moderate to severe chronic illnesses (according to the International Classification of Diseases-9; see ^43^). For BASE-II, we computed a morbidity index largely based on the categories of the Charlson index ^44^, which is a weighted sum of moderate to severe, mostly chronic physical illnesses, see ^42^.

##### Lung capacity

Forced expiratory volume in one second (FEV1) was used as an overall indicator of lung function. We only analyzed spirometry measurements (using EasyOne Spirometer; ndd Medical Technologies) with sufficient measurement quality, fully in line with standard procedures following the guidelines of the American Thoracic Society ^45^. *Subjective health* was assessed using a single item measure asking individuals how they would rate their overall health on a scale from 1 (very bad) to 5 (very good).

##### Alternative age biomarkers

To ensure that our results were not confounded by alternative age biomarkers that are known to be associated with biological age ^6,7^, we made use of the rich interdisciplinary data in BASE-II that allowed examining telomere length, DNA methylation age, skin age, and subjective age. The measurement of *Relative leukocyte telomere length (rLTL)* is described in detail in Meyer et al.^13^. Briefly, genomic DNA was extracted from EDTA blood using the LGC ‘Plus XL manual kit’ (LGC, Germany, Berlin). rLTL was measured using a modified quantitative PCR protocol originally described by Cawthon and colleagues (2002). All samples were measured in triplicate and their mean was used for further analysis when the ct values of both PCRs (telomere PCR and single copy gene [36B4] PCR) showed a variation coefficient < 2%. The rLTL was subsequently calculated according to Pfaffl and colleagues (2001). Pooled DNA from 10 randomly selected subjects was used as the reference (rLTL = 1).

##### DNA methylation age

Genomic DNA was extracted with the LGC “Plus XL manual kit” (LGC) from EDTA blood and used for methylation analysis. An adapted protocol from Vidal-Bralo and colleagues ^18^ was used to measure the fraction of the methylated cysteine bases. DNA methylation age was calculated based on seven CpG sites (for further details, please see ^17^). *Skin aging* was estimated by quantifying lentigines on both hands from hand photographs taken of each BASE-II participant. Photos were taken at the baseline assessment and examined independently by three investigators in three rating sessions. The amount and size of lentigines on the back of the hands was quantified using a four-level categorical score (ranging from 0 [no or very few lentigines] to 3 [very abundant presence of lentigines on both hands]). The resulting lentigines score per BASE-II participant represents the weighted average of the score assigned to each photograph by the three reviewers. *Subjective age* was assessed by asking participants how old they felt in years ^46^. In line with previous research ^47,48^, we calculated proportional discrepancy scores by subtracting participants’ subjective age from their chronological age and then dividing by chronological age. Positive scores indicate a younger subjective age. Following usual practice ^47,49^, proportional discrepancy scores three standard deviations above or below the mean were considered outliers and replaced with a score equivalent to the mean plus or minus three standard deviations, respectively; this was necessary for one participant only.

For BASE, information about *mortality* status and date of death for deceased participants has been updated regularly from the Berlin city registry since study inception in 1990. Our information on death makes use of data from a November 2016 update, when, of the 516-sample, 514 participants were known to have died and 2 were still alive.

### Statistical Procedure

In a preliminary step, we identified a total of 33 blood laboratory parameters that had been available in both studies for the majority of participants and that had been assessed in highly comparable ways. With these variables, we estimated a series of separate Cox proportional hazards regression models ^31^ to identify those parameters that have been predictive of mortality hazards. In a second step, we used the parameters identified in the preceding step as being mortality-relevant to calculate the biological age composite following procedures described in ^27^. Specifically, we regressed *m* number of biomarkers on age and calculated biological age of individual *i* as:

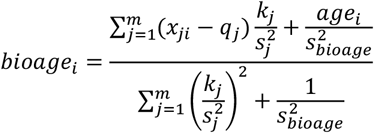

where *q* (intercept), *k* (slope) and *s* (root mean squared error) are parameters from these regressions, and age is the chronological age of a participant. With the biological age composite now being compiled for BASE, we conducted descriptive analyses linking biological age with chronological age and examining the predictive effects of the biological age composite for mortality hazards. In a third step, we used regression analyses to examine the predictive role of biological age for individual differences in socio-demographic and physical health variables. In a fourth step, we used *the exact same* biomarkers identified in BASE as mortality-relevant to compute the biological age composite in BASE-II. In a fifth step, we repeated the analyses noted to examine similarities and differences in the nature and correlates of biological age between BASE and BASE-II. Finally, we examined the predictive validity of biological age for several health measures while at the same time accounting for alternative age biomarkers.

## Data Availability

The data can be requested from the steering committee of the Berlin Aging Study and the Berlin Aging Study II. Further details about the procedure can be obtained at https://www.base-berlin.mpg.de/de and www.base2.mpg.de.

## Acknowledgements

This article reports data from the Berlin Aging Study (BASE; www.base-berlin.mpg.de). The BASE was initiated by the late Paul B. Baltes, in collaboration with Hanfried Helmchen, psychiatry; Elisabeth Steinhagen-Thiessen, internal medicine and geriatrics; and Karl Ulrich Mayer, sociology (amongst the post-docs were Ulman Lindenberger and Gert G. Wagner). Further details about BASE-II can be obtained at https://www.base2.mpg.de/en.

## Statement of Ethics

The research was conducted ethically in accordance with the <u>World Medical Association Declaration of Helsinki.</u> Subjects gave their written informed consent and that the study protocol was approved by the ethics committee of the Max Planck Institute for Human Development, Berlin, Germany, Charité-Universitaetsmedizin Berlin, and DIW BERLIN.

## Author Contributions

Design of the work: J.D., G.H., I.D., D.G.

Data acquisition: J.D., S.D., I.D., U.L., G.G.W., E.S-T., L.B., C. L., D.G., V.M.V.

Data analysis: J.D., G.H., D.G.

Writing of paper: J.D., I.D., D.G., G.H.

All authors critically and substantively revised the manuscript.

## Supplementary Material

**Table A.1.** Hazard Ratios for Mortality by Variables Defining Biological Age in the Berlin Aging Study

|  | Hazard Ratio | [95% CI] | *N* |
| --- | --- | --- | --- |
| Zinc | 0.858* | 0.779–0.945 | 489 |
| Sodium | 0.857* | 0.781–0.941 | 505 |
| Chloride | 0.902* | 0.824–0.986 | 502 |
| Uric acid | 1.235* | 1.120–1.363 | 504 |
| Albumin | 0.893* | 0.819–0.974 | 505 |
| Alpha1 globulin | 1.139* | 1.050–1.236 | 498 |
| Alpha2 globulin | 1.138* | 1.037–1.250 | 498 |
| HbA1c | 1.243* | 1.124–1.376 | 451 |
| Hemoglobin | 0.856* | 0.765–0.959 | 494 |
| Leukocytes | 1.792* | 1.374–2.336 | 494 |
| Lymphocytes | 0.543* | 0.410–0.720 | 477 |
| Creatinine normed | 0.843* | 0.735–0.967 | 507 |

*Note.* Each row reports hazard ratios from separate models that included a given marker (z-standardized) along with chronological age, sex, and education. The only exceptions are coefficients for leukocytes and lymphocytes that were obtained from a conjoint model including both markers (in univariate analyses, the predictive effects of lymphocytes did not reach conventional levels of statistical significance). For parsimony, the predictive effects of chronological age, sex, and SES are not reported.
\* $p < .05$ .

**Table A.2.** Hazard Ratios for Mortality by Variables not used to define Biological Age in the Berlin Aging Study

|  | Hazard Ratio | [95% CI] | N |
| --- | --- | --- | --- |
| Lipase | 0.999 | 0.909–1.099 | 502 |
| Potassium | 1.012 | 0.920–1.112 | 504 |
| Calcium | 0.953 | 0.865–1.050 | 503 |
| Phosphate | 1.015 | 0.922–1.119 | 503 |
| Protein | 0.955 | 0.872–1.045 | 505 |
| Bilirubin | 0.982 | 0.884–1.090 | 504 |
| Cholesterol | 0.943 | 0.853–1.042 | 505 |
| SGOT | 0.956 | 0.867–1.054 | 506 |
| SGPT | 0.962 | 0.871–1.062 | 506 |
| SGGT | 1.038 | 0.975–1.105 | 506 |
| Alkaline Phosphatase | 1.017 | 0.934–1.107 | 507 |
| Beta globulin | 0.981 | 0.900–1.069 | 498 |
| Gamma globulin | 1.073 | 0.980–1.176 | 498 |
| Erythrocytes | 0.911 | 0.813–1.021 | 494 |
| Mean Corpuscular Hemoglobin | 0.939 | 0.844–1.044 | 494 |
| Mean Corpuscular Hemoglobin Concentration | 0.898 | 0.809–0.997 | 494 |
| Mean Corpuscular Volume | 0.989 | 0.889–1.099 | 494 |
| Thrombocytes | 1.075 | 0.972–1.189 | 493 |
| Quick | 0.958 | 0.872–1.052 | 489 |
| PTT | 0.975 | 0.900–1.056 | 487 |
| Fibrinogen | 1.087 | 0.996–1.187 | 468 |

*Note.* Each row reports hazard ratios from separate models that included a given marker (z-standardized) along with chronological age, sex, and education. For parsimony, the predictive effects of chronological age, sex, and SES are not reported.
\* $p < .05$ .

**Table A.3.**
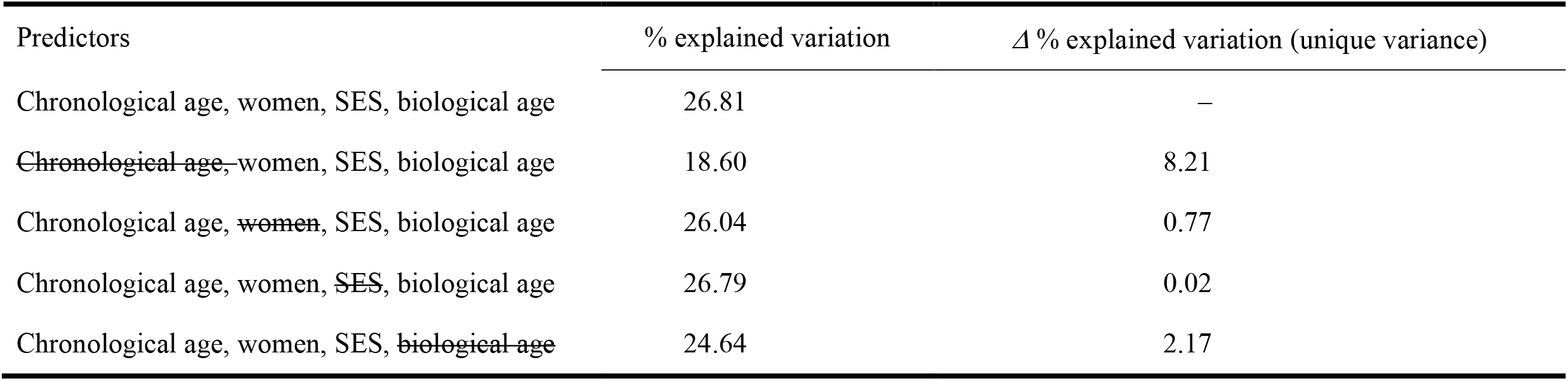
Testing the unique predictive effects of biological age vis-a-vis those of chronological age, sex, and SES.

**Table A.4.** Intercorrelations among the Alternative Age Markers in the Berlin Aging Study II

|  | Intercorrelations |  |  |  |  |
| --- | --- | --- | --- | --- | --- |
|  | 1 | 2 | 3 | 4 | 5 |
| (1) Telomere length (0.17–1.93) | 1 | −.01 | .01 | −.02 | .00 |
| (2) DNA methylation age – chronological age (−22.93–26.61) | −.01 | 1 | .23* | .03 | .14* |
| (3) Biological age – chronological age (−28.52–22.77) | .02 | .04 | 1 | .10* | .19* |
| (4) Skin age (0–3) | −.02 | −.04 | .02 | 1 | .09* |
| (5) Proportional subjective age (−0.66–0.15) | .05 | −.04 | −.04 | .01 | 1 |
| *N* | 1,460 | 1,395 | 1,517 | 1,240 | 1,382 |
| *M* | 1.14 | 0.02 | 0 | 1.72 | −0.12 |
| *SD* | 0.23 | 6.91 | 6.10 | 0.90 | 0.09 |

*Note.* Raw intercorrelations between telomere length, DNA methylation age, biological age, skin age, and subjective age are reported above the diagonal, intercorrelations that are residualized for age are reported below the diagonal.
\* $p < .05$ .

## References

1. Baltes, P. B., Lindenberger, U. & Staudinger, U. M. Life Span Theory in Developmental Psychology. in Handbook of Child Psychology (John Wiley & Sons, Inc., 2006). doi:10.1002/9780470147658.chpsy0111.

2. Anne Nelson, E. & Dannefer, D. Aged Heterogeneity: Fact or Fiction? The Fate of Diversity in Gerontological Research. Gerontologist 32, 17–23 (1992).

3. Jylhävä, J., Pedersen, N. L. & Hägg, S. Biological Age Predictors. EBioMedicine vol. 21 29–36 (2017).

4. Butler, R. N. et al. Biomarkers of Aging: From Primitive Organisms to Humans. (2004).

5. Johnson, T. E. Recent results: Biomarkers of aging. Experimental Gerontology 41, 1243–1246 (2006).

6. Jylhävä, J., Pedersen, N. L. & Hägg, S. Biological Age Predictors. EBioMedicine vol. 21 29–36 (2017).

7. Butler, R. N. et al. Biomarkers of aging: from primitive organisms to humans. The journals of gerontology. Series A, Biological sciences and medical sciences 59, 560–567 (2004).

8. Bekaert, S., de Meyer, T. & van Oostveldt, P. Telomere Attrition as Ageing Biomarker. ANTICANCER RESEARCH 25, 3011–3022 (2005).

9. Rode, L., Bojesen, S. E., Weischer, M., Vestbo, J. & Nordestgaard, B. G. Short telomere length, lung function and chronic obstructive pulmonary disease in 46 396 individuals. Thorax 68, 429–435 (2013).

10. Weischer, M. et al. Short telomere length, myocardial infarction, ischemic heart disease, and early death. Arteriosclerosis, Thrombosis, and Vascular Biology 32, 822–829 (2012).

11. Zee, R. Y. L., Castonguay, A. J., Barton, N. S., Germer, S. & Martin, M. Mean leukocyte telomere length shortening and type 2 diabetes mellitus: a case-control study. Translational Research 155, 166–169 (2010).

12. Saßenroth, D. et al. Sports and Exercise at Different Ages and Leukocyte Telomere Length in Later Life--Data from the Berlin Aging Study II (BASE-II). PloS one 10, (2015).

13. Meyer, A., Salewsky, B., Buchmann, N., Steinhagen-Thiessen, E. & Demuth, I. Relative Leukocyte Telomere Length, Hematological Parameters and Anemia - Data from the Berlin Aging Study II (BASE-II). Gerontology 62, 330–336 (2016).

14. Meyer, A. et al. Leukocyte telomere length is related to appendicular lean mass: cross-sectional data from the Berlin Aging Study II (BASE-II). The American journal of clinical nutrition 103, 178–183 (2016).

15. Hannum, G. et al. Genome-wide Methylation Profiles Reveal Quantitative Views of Human Aging Rates. Molecular cell 49, 359 (2013).

16. Horvath, S. DNA methylation age of human tissues and cell types. Genome Biology 14, 1–20 (2013).

17. Vetter, V. M. et al. Epigenetic Clock and Relative Telomere Length Represent Largely Different Aspects of Aging in the Berlin Aging Study II (BASE-II). The journals of gerontology. Series A, Biological sciences and medical sciences 74, 27–32 (2019).

18. Vidal-Bralo, L., Lopez-Golan, Y. & Gonzalez, A. Simplified Assay for Epigenetic Age Estimation in Whole Blood of Adults. Frontiers in genetics 7, (2016).

19. Banszerus, V. L., Vetter, V. M., Salewsky, B., König, M. & Demuth, I. Exploring the Relationship of Relative Telomere Length and the Epigenetic Clock in the LipidCardio Cohort. International journal of molecular sciences 20, (2019).

20. Belsky, D. W. et al. Eleven Telomere, Epigenetic Clock, and Biomarker-Composite Quantifications of Biological Aging: Do They Measure the Same Thing? American journal of epidemiology 187, 1220–1230 (2018).

21. Breitling, L. P. et al. Frailty is associated with the epigenetic clock but not with telomere length in a German cohort. Clinical epigenetics 8, 1–8 (2016).

22. Chen, B. H. et al. Leukocyte telomere length, T cell composition and DNA methylation age. Aging (Albany NY) 9, 1983 (2017).

23. Marioni, R. E. et al. The epigenetic clock and telomere length are independently associated with chronological age and mortality. International journal of epidemiology 45, 424–432 (2018).

24. Kwon, D. & Belsky, D. W. A toolkit for quantification of biological age from blood chemistry and organ function test data: BioAge. GeroScience 43, 2795 (2021).

25. Levine, M. E. Modeling the rate of senescence: can estimated biological age predict mortality more accurately than chronological age? The journals of gerontology. Series A, Biological sciences and medical sciences 68, 667–674 (2013).

26. Klemera, P. & Doubal, S. A new approach to the concept and computation of biological age. Mechanisms of ageing and development 127, 240–248 (2006).

27. Belsky, D. W. et al. Quantification of biological aging in young adults. Proceedings of the National Academy of Sciences of the United States of America 112, E4104–E4110 (2015).

28. Sebastiani, P. et al. Biomarker signatures of aging. Aging cell 16, 329–338 (2017).

29. Zhang, Q. et al. Improved precision of epigenetic clock estimates across tissues and its implication for biological ageing. Genome Medicine 11, (2019).

30. Alpert, A. et al. A clinically meaningful metric of immune age derived from high-dimensional longitudinal monitoring. Nature Medicine 2019 25:3 25, 487–495 (2019).

31. Cox, D. R. Regression Models and Life-Tables. Journal of the Royal Statistical Society: Series B (Methodological) 34, 187–202 (1972).

32. Bertram, L. et al. Cohort profile: The Berlin Aging Study II (BASE-II). International journal of epidemiology 43, 703–712 (2014).

33. Gerstorf, D. et al. Secular changes in late-life cognition and well-being: Towards a long bright future with a short brisk ending? Psychology and Aging 30, 301–310 (2015).

34. Hülür, G. et al. Cohort Differences in Psychosocial Function over 20 Years: Current Older Adults Feel Less Lonely and Less Dependent on External Circumstances. Gerontology 62, 354–361 (2016).

35. König, M. et al. Historical trends in modifiable indicators of cardiovascular health and self-rated health among older adults: Cohort differences over 20 years between the Berlin Aging Study (BASE) and the Berlin Aging Study II (BASE-II). PloS one 13, (2018).

36. Deelen, J. et al. A metabolic profile of all-cause mortality risk identified in an observational study of 44,168 individuals. Nature Communications 2019 10:1 10, 1–8 (2019).

37. Ashiqur Rahman, S. et al. Deep learning for biological age estimation. Briefings in Bioinformatics 22, 1767–1781 (2021).

38. Schrempft, S. et al. Associations between life course socioeconomic conditions and the Pace of Aging. The Journals of Gerontology. Series A, Biological Sciences and Medical Sciences (2021) doi:10.1093/gerona/glab383/6482783.

39. Vidal-Pineiro, D. et al. Individual variations in ‘brain age’ relate to early-life factors more than to longitudinal brain change. eLife 10, (2021).

40. Mayer, K. U., Maas, I. & Wagner, M. Socioeconomic conditions and social inequalities in old age. in The Berlin Aging Study: Aging from 70 to 100 (eds. Baltes, P. B. & Mayer, K. U.) 227–255 (Cambridge University Press, 1999).

41. Gerstorf, D., Ram, N., Lindenberger, U. & Smith, J. Age and time-to-death trajectories of change in indicators of cognitive, sensory, physical, health, social, and self-related functions. Developmental Psychology 49, 1805–1821 (2013).

42. Drewelies, J. et al. Location, Location, Location: The Role of Objective Neighborhood Characteristics for Perceptions of Control. Gerontology 1–10 (2021) doi:10.1159/000515634.

43. Steinhagen-Thiessen, E. & Borchelt, M. Morbidity, medication, and functional limitations in very old age. in The Berlin Aging Study: Aging from 70 to 100 (eds. Baltes, P. B. & Mayer, K. U.) 131–166 (Cambridge University Press, 1999).

44. Charlson, M. E., Pompei, P., Ales, K. L. & MacKenzie, C. R. A new method of classifying prognostic comorbidity in longitudinal studies: development and validation. Journal of chronic diseases 40, 373–383 (1987).

45. Hill, K. et al. Prevalence and underdiagnosis of chronic obstructive pulmonary disease among patients at risk in primary care. CMAJ : Canadian Medical Association journal = journal de l’Association medicale canadienne 182, 673–678 (2010).

46. Notthoff, N. et al. Feeling older, walking slower—but only if someone’s watching. Subjective age is associated with walking speed in the laboratory, but not in real life. European Journal of Ageing 15, 425–433 (2018).

47. Stephan, Y., Sutin, A. R. & Terracciano, A. Subjective Age and Personality Development: A 10-Year Study. Journal of Personality 83, 142–154 (2015).

48. Rubin, D. C. & Berntsen, D. People over forty feel 20% younger than their age: Subjective age across the lifespan. Psychonomic Bulletin & Review 2006 13:5 13, 776–780 (2006).

49. Weiss, D. & Lang, F. R. “They” are old but “I” feel younger: Age-group dissociation as a self-protective strategy in old age. Psychology and Aging 27, 153–163 (2012).

